# Automatic Detection of COVID-19 Infection from Chest X-ray using Deep Learning

**DOI:** 10.1101/2020.05.10.20097063

**Authors:** Kishore Medhi, Md. Jamil, Md. Iftekhar Hussain

## Abstract

COVID-19 infection has created a panic across the globe in recent times. Early detection of COVID-19 infection can save many lives in the prevailing situation. This virus affects the respiratory system of a person and creates white patchy shadows in the lungs. Deep learning is one of the most effective Artificial Intelligence techniques to analyse chest X-ray images for efficient and reliable COVID-19 screening. In this paper, we have proposed a Deep Convolutional Neural Network method for fast and dependable identification of COVID-19 infection cases from the patient chest X-ray images. To validate the performance of the proposed system, chest X-ray images of more than 150 confirmed COVID-19 patients from the Kaggle data repository are used in the experimentation. The results show that the proposed system identifies the cases with an accuracy of 93%.

## 1. Introduction

The COVID-19 commonly known as the novel coronavirus has become a pandemic disease causing a total of 2,01,502 deaths till 26^th^ April 2020 [16]. Since the first reporting in the Wuhan city of China in December 2019, it has spread to most of the countries of the world. Subsequently, the World Health Organization (WHO) has declared this ongoing outbreak as a global public health emergency [1]. The total number of confirmed and fatal COVID-19 cases in some of the highly affected countries are shown in Fig. 1. COVID-19 mainly affects the respiratory system of human and damages the alveoli of lungs. Although COVID-19 infection cases are mild in general, it causes severe disease with 2% fatality rate [17].

**Fig. 1:**
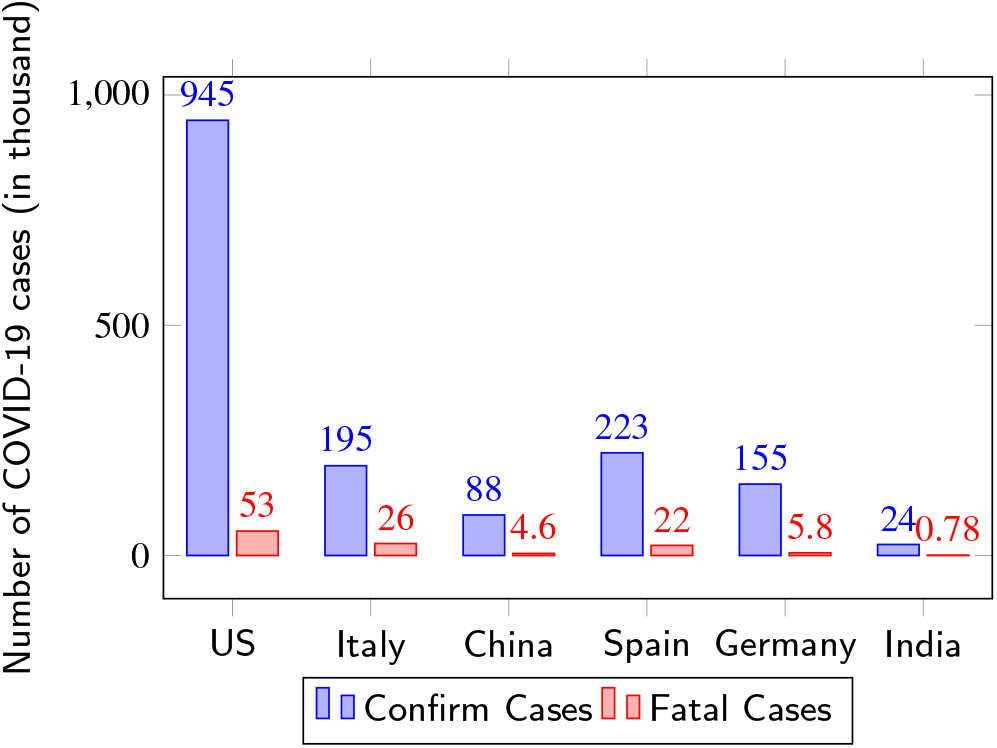
Number of confirmed and fatal cases in US, Italy, China, Spain, Germany, and India as on 26*^th^* April,2020

This COVID-19 virus transmits from person to person and creates pneumonia in the human body. The clinical symptoms of COVID-19 infection includes respiratory tract illness, severe viral pneumonia with respiratory failure [18]. F. Zhou et al. [18] made a survey of 813 adult patients who were hospitalised in Wuhan Pulmonary Hospital and found that most of the patients who died were male. Different diseases were present in nearly half of the cases, hypertension being the most common, followed by coronary heart disease and diabetes. The regular symptoms on hospital admission were cough and high fever, followed by sputum production and fatigue. In a serious situation, the infection can cause pneumonia, multi-organ failure, and even death [8].

The infection of COVID-19 virus can easily be identified by using the X-ray of chest. A chest X-ray shows multiple white patchy shadows in both the lungs of a COVID-19 affected person [17]. Artificial Intelligence (AI) with image processing could be an efficient and accurate technique to differentiate this normal and affected chest X-ray images with good accuracy. It has the potential to improve the speed of COVID-19 case identification and reduce its spread particularly in resource-constraint environments where expert radiologist and high-end medical equipment are not available for early diagnosis and management of patients [10].

Deep Convolutional Neural Network is an important AI tool which is used for detection of diseases from the medical images ([7], [9], [11]). Lopes et al. [7] identified tuberculosis from chest X-ray images using the CNN technique with an accuracy of more than 92%. Sergio et al. [9] and Saman et al. [11] detected Alzheimer and Brain tumor from MRI images using the same technique. Similarly, Ali et al. [8] and Linda et al. [15] developed a ResNet architecture and a CNN architecture respectively to detect COVID-19 infection from 76 and 50 confirmed chest X-ray images respectively. In both of these experiments, they directly applied the feature extraction and classification processes to the raw images without any segmentation and removal of the artifacts. However both of these steps are very important to reduce the false COVID-19 alarm. Nevertheless, all these contributions provide motivation to use AI techniques in identifying diseases accurately with lesser time and without any human expert intervention.

In this research, we have used CNN to identify the presence of COVID-19 infection from the chest X-ray image of a patient. To improve the accuracy level further, we have used a Gaussian filter to reduce the artifact, and a segmentation technique based on the gray level thresholding to extract the important portion from the X-ray images. To conduct the experiment, we have used a chest X-ray image dataset collected from Kaggle [6]. Performance of the proposed system is measured and compared with some relevant schemes in terms of accuracy, false positives, and false negatives. Our proposed scheme is seen to outperform those in all the aspects.

The rest of the paper is organized as follows. Section II describes the details about the dataset used in this study. Analysis of various clinical symptoms is presented in Section 3. Section 4 describes the details of the proposed method. At last, result analysis and conclusion is presented in Section 5 and 6 respectively.

## 2. Dataset

In this research, we have used two different datasets: one for statistical analysis of symptoms [13] and the other for chest X-ray identification [6]. Both the datasets are collected from the Kaggle data repository. The first dataset was generated by John Hopkins University [14] which collected the daily information from the COVID-19 affected cases, deaths and recovery statistics of Wuhan City. This dataset contains more than 14000 patients information with 44 different attributes including age, gender, date of onset, date of confirmation, travel history, symptoms, chronic diseases, etc. The second dataset contains chest X-ray images along with clinical symptoms of more than 150 different COVID-19 patients collected from Wuhan city.

## 3. Analysis of COVID-19 clinical symptoms

The initial symptoms of COVID-19 includes fever, cough, dyspnea, myalgia or fatigue, headache, sputum production, diarrhea, hemoptysis, etc. [5]. To explore the prominent symptoms associated with this disease, we have analysed an COVID-19 dataset collected from kaggle. Detail description of the dataset is available in the Section 2. The analysis revealed that 12% of the total patients died with an average age of 65.4 years. In that dataset, 59% of the total COVID-19 patients were male and the remaining 41% were female. Fever and cough were the most prominent characteristics of COVID-19 patients being available with 95% and 71% cases respectively. It was also observed that 95% of patients who died had fever whereas 80% had pneumonia. As shown in Table. 2, the average time from illness onset to hospital admission was high in case of death patient compared to discharged patient.

**Table 1.** Clinical symptoms of COVID-19 patients

| Feature | Total Patients | Death | Discharge |
| --- | --- | --- | --- |
| Age (in year) | (2-89) | (44-89) | (7-73) |
| Male | 59% | 59% | 61% |
| Female | 41% | 41% | 39% |
| Fever | 95% | 95% | 92% |
| Caugh | 71% | 33% | 66% |
| Pneumonia | 75% | 80% | 50% |
| Headache | 75% | 60% | 55% |
| Diarrhea | 10% | 5% | 3% |

**Table 2.**
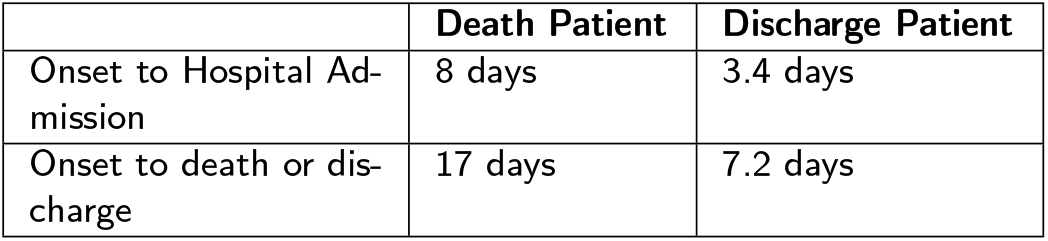
Analysis of average time from the date of illness onset to hospital admission, discharge, and death

In addition to these clinical symptoms, COVID-19 also shows its effect on lungs. A detail comparison of normal and COVID-19 patient lungs has been shown in Fig. 2. It clearly shows the white patchy ground-glass shadows present in the lungs of COVID-19 patient. H. Chen et al. [1] performed a study on lungs of nine different corona patients. They collected the different Chest X-ray and CT scan images from these patients. The study found that out of nine, eight patients had white patchy ground glass shadow in lungs.

**Fig. 2:**
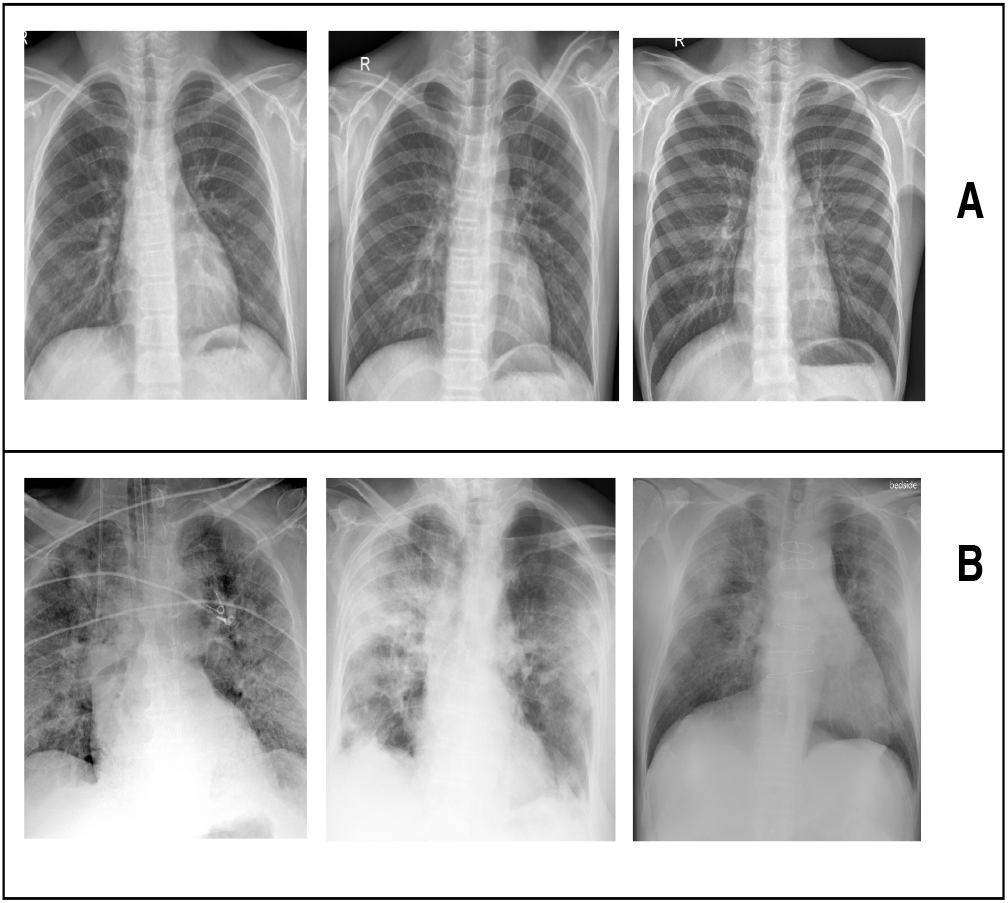
Comparison of different chest X-ray of normal & COVID-19 patient’s lungs. SET A are normal and SET B are COVID-19 lungs

## 4. The Proposed Method

The automatic COVID-19 detection based on the chest X-ray image is a complicated image identification problem due to varying image quality across equipments used and patients. As shown in Fig. 3, the typical workflow of our proposed system mainly consists of five major steps: i) data acquisition, ii) pre-processing, iii) segmentation, iv) feature extraction, and v) classification model. These steps are explained in the following subsections.

**Fig. 3:**
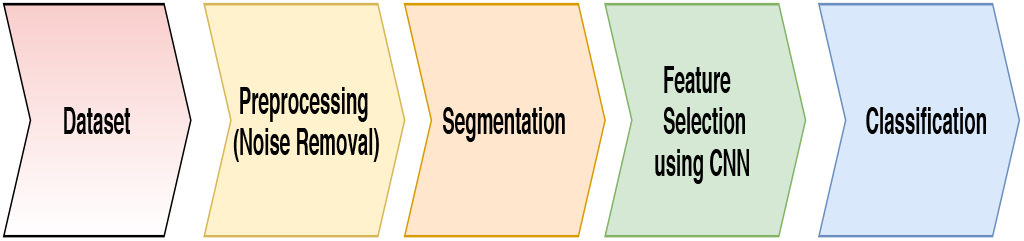
Flow diagram of the proposed system

### 4.1. Data Acquisition

To carry out the experiment, we have collected the chest X-ray image dataset from kaggle. This dataset was generated from the admitted patients in designated hospitals located in Wuhan city of China from Dec 16, 2019 to Jan 2, 2020.

### 4.2. Pre-processing

Pre-processing is an essential step in classification process to improve the system accuracy. In pre-processing, we have removed the noise present in the images. In medical images, Gaussian noise is common due to thermal fluctuations of electronic components [4]. In X-ray images, the amount of noise present is dependent on the sensitivity of the receptor [3]. In order to reduce this noise, we have used the 2D Gaussian filter. A 2D Gaussian filter can be mathematically expressed as follows:

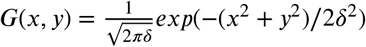

Here, *δ*^2^ is the variance and size of the filter kernal *l* (−*l* <= *x, y* <= *l*) [2]. By applying this filter, the Signal to Noise Ratio (SNR) of the images have been enhanced. In Fig. 4, we have shown the filtered and original X-ray images with 10dB and 4dB SNR values respectively.

**Fig-4:**
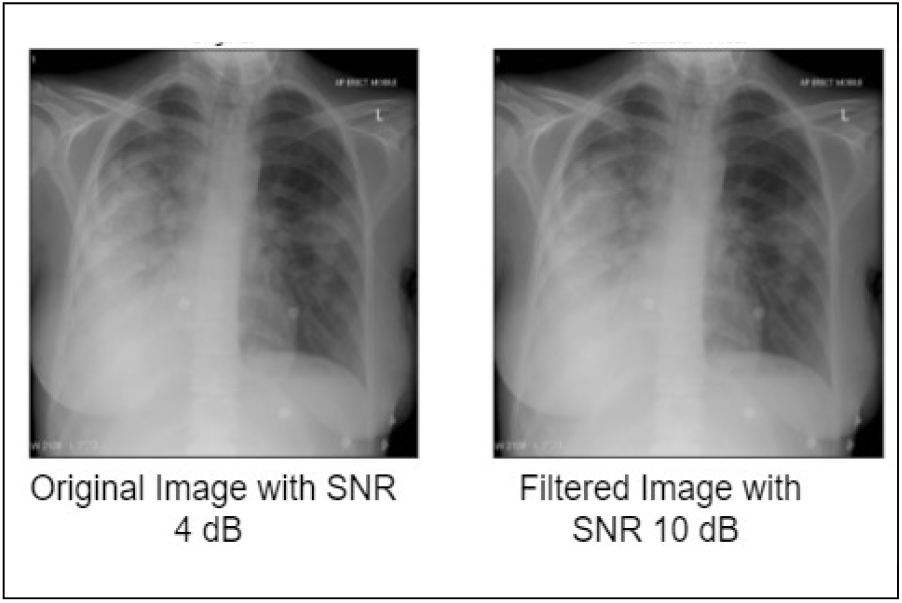
Removal of noise from chest X-ray image using Gaussian filter. First image is the original image with 4 dB SNR value and second image is the filtered image with 10dB SNR value.

### 4.3. Segmentation

Segmentation is the method of dividing an image into some sections with related properties such as gray level, color, brightness, and contrast. Proper segmentation of medical images is an essential step in disease identification by identifying the region of interest, locating the tumor, measuring the tissue volume, etc. In this research, we have segmented the chest X-ray images based on the gray level thresholding as the lungs of a COVID-19 patient contains many white patches. All the gray regions are separated from others by converting gray pixels into white pixels and remaining into black pixels. In this way, one can easily differentiate the normal and abnormal chest X-ray images. Segmentation of normal and COVID-19 patient lungs images are shown in Fig.5. Here, set A and set B show normal lungs and COVID-19 lungs respectively. It can clearly be observed that the COVID-19 lungs has less white pixels compared to normal one. Mathematically the gray level thresholding can be expressed as shown in the following equation.

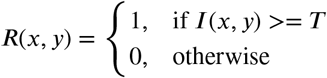

**Fig. 5:**
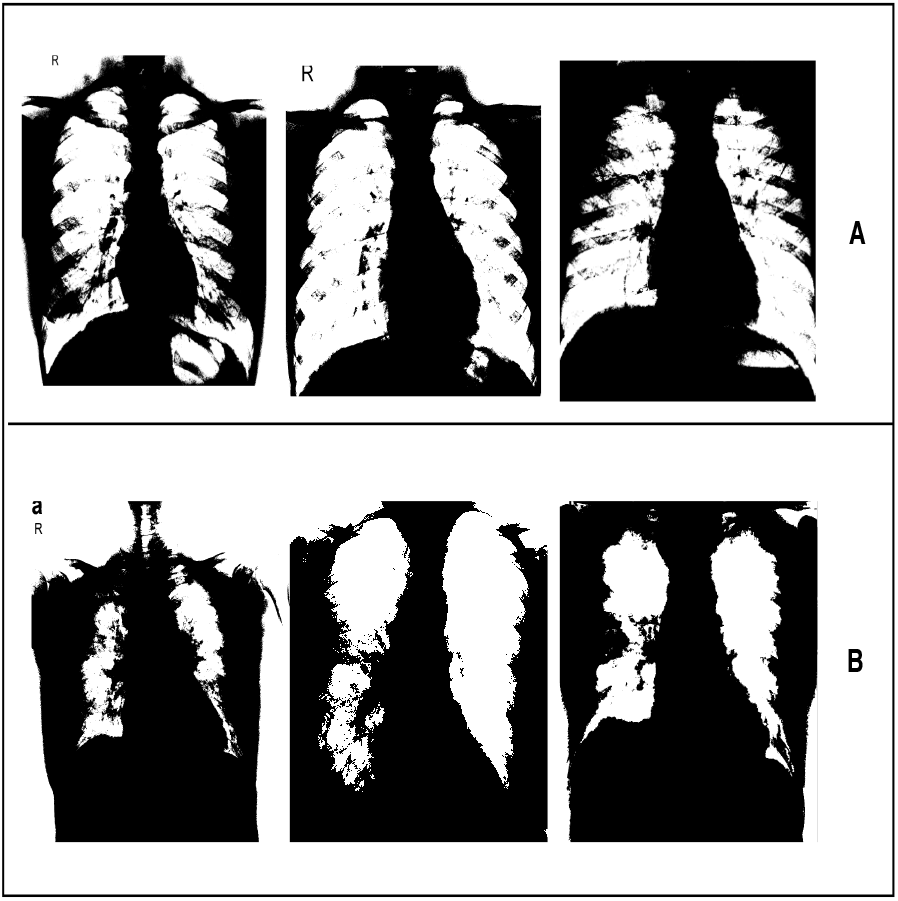
Segmentation of normal and COVID-19 chest X-ray image. SET A are normal and SET B are COVID-19 lungs images after segmentation.

Here, I(x, y) and R(x, y) are the input image and resulting image pixels respectively and T is the threshold value. Proper threshold selection is an important task in threshold-based segmentation techniques to improve segmentation efficiency. Various researchers use the histogram of an image to determine the threshold value based on the pixel value with minimum frequency [12]. Histogram of chest X-ray image is shown in Fig. 6, where image A and B are of normal and COVID-19 affected X-ray images respectively. From this figure it can be observed that the pixel value with minimum frequency lies between 25 to 30 for both the images. Hence we have set the threshold value as 30.

**Fig. 6:**
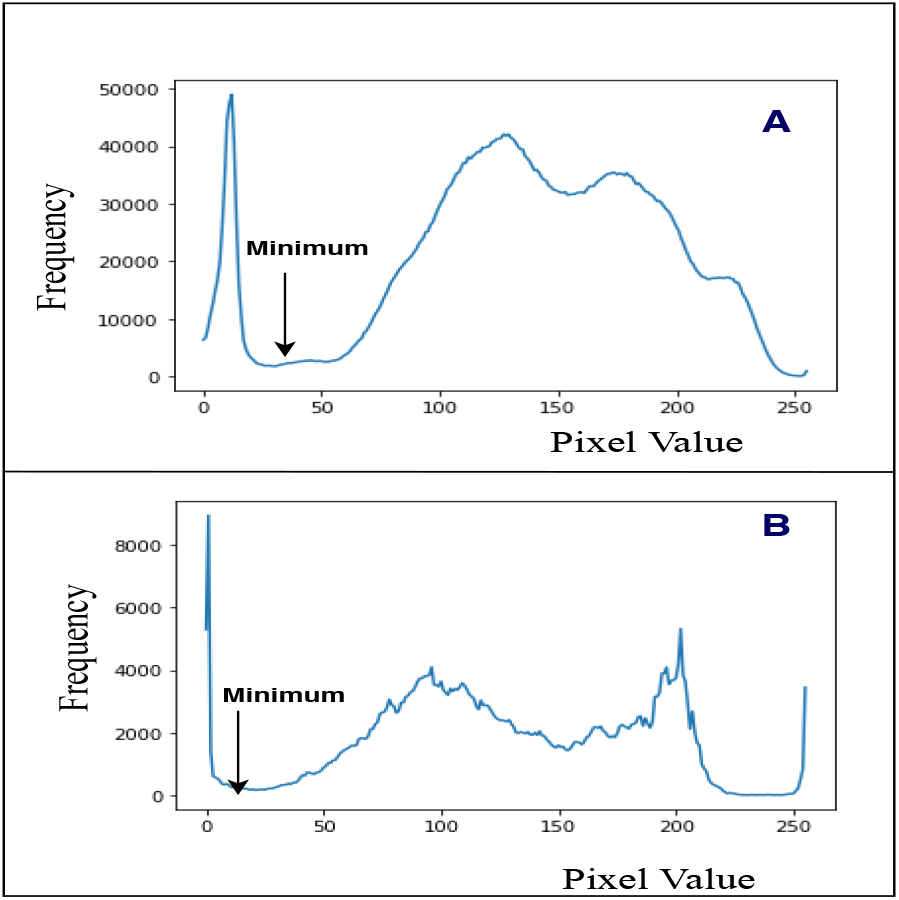
Histogram of the chest X-ray image. Here, image A and B is the histogram of normal and COVID-19 infected X-ray image respectively.

### 4.4. Feature Extraction

A multi-layer CNN architecture is developed to extract the important features from the X-ray images for automatic identification of COVID-19 infection which is shown in Fig. 7. Convolution layer produces some high-level complex features which are reduced by the pooling layers. In this progressive manner, CNN can learn hierarchical features layer by layer automatically. The developed CNN architecture consists of four convolutional layers, four maxpooling layers and we have used ReLU activation function. During convolution we have used a kernal size of 3*x*3 and 0 stride size during pooling. After final convolution and maxpooling we have finally received 22 features value as output.

**Fig. 7:**
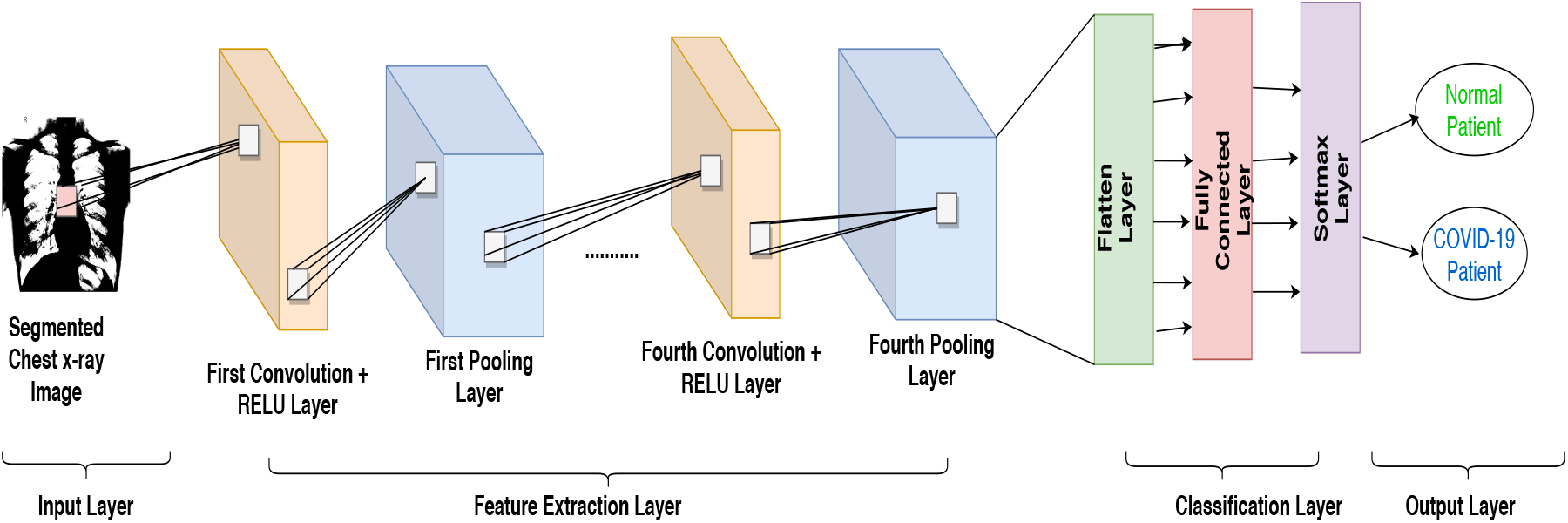
Architectire of the proposed CNN model

### 4.5. Classification Model

The fully connected layer is an essential component of CNN; it helps in the classification of an object. It takes input from the output of the previous layer i.e., from the last feature extraction layer. The flatten layer turns the feature matrix into a single vector and passes it to the first fully connected layer. These feature values are multiplied by weight vectors and passed to the softmax activation function just like a traditional artificial neural network. To learn the feature more accurately and to reduce the error, weights of the fully connected layers are updated by using the back-propagation algorithm. The neuron of the output layer gives the classification level of each input image. In our classification model, there are two fully connected layers and one softmax activation layer. Each fully connected layer has 128 numbers of neurons whereas the output layer has only one.

**Table 3.** List of parameters for the proposed CNN architecture

| Layers | Kernal Size, no. of filters | Maxpool, Stride | Output size |
| --- | --- | --- | --- |
| Convolution1 | (3×3), filter 65025 |  | 65536 |
| Maxpool1 |  | (3×3), stride 0 | 7281 |
| Convolution2 | (3×3), filter 7112 |  | 7281 |
| Maxpool2 |  | (3×3), stride 0 | 809 |
| Convolution3 | (3×3), filter 754 |  | 809 |
| Maxpool3 |  | (3×3), stride 0 | 90 |
| Convolution4 | (3×3), filter 73 |  | 90 |
| Maxpool4 |  | (2×2), stride 0 | 22 |

## 5. Result & Analysis

In this research to identify the presence of Covid-19 infection, we have used chest x-ray images. The identification process is based on the presence of a white patchy shadow in the lungs. For proper detection, we have pre-processed all the images by segmenting the important lungs section from the others. Before segmentation, we have cleared all the artifacts using the Gaussian filter. In this experiment, a threshold-based segmentation is done to separate gray pixels from the others. During the segmentation, we have set the threshold value into 30. In noise removal step, the image quality has increased from SNR 4 dB to 10 dB. The output of segmentation has shown in Fig. 5, where we have clearly separated the gray pixels from the white pixels. From the segmentation results, it is also clear that the number of gray pixels in Covid-19 affected lungs is very less compared to the normal lungs. After segmentation, features were extracted from the segmented images using the above mentioned CNN architecture and finally all the extracted features were fed into the softmax classifier for the detection. During the experiment, the dataset was divided into two parts where 80% was kept for training and the remaining 20% was used for testing. The experimental results of our proposed system during training and testing has shown in Table. 4. To analyse the performance of our proposed system, we have used a confusion matrix as shown in Fig. 9. It measures three different parameters: accuracy, false positive, and false negative using the equations 1, 2, and 3 respectively.

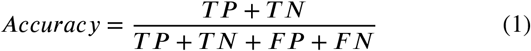

**Fig. 8:**
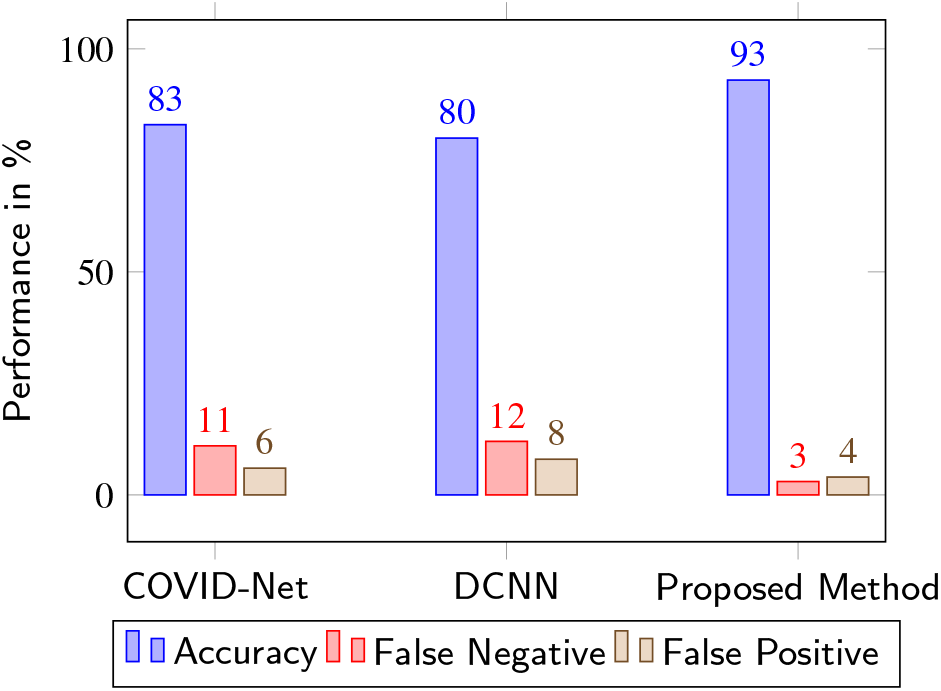
Performance comparison of various methods with the proposed scheme

**Fig. 9:**
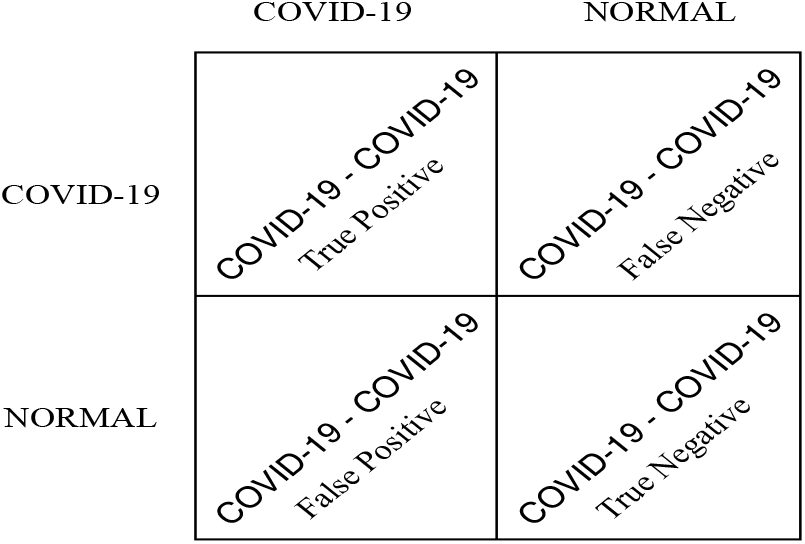
Confusion matrix to measure accuracy, false negative, and false positive

**Table 4.** Performance analysis of the proposed system during training and testing

| Parameter | Training Result (in %) | Testing Result (in %) |
| --- | --- | --- |
| Accuracy | 100 | 93 |
| False Positive | 0 | 4 |
| False Negative | 0 | 3 |

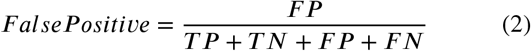

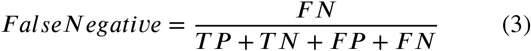

Here TP, TN, FP, and FN represent the number of True Positive, True Negative, False Positive, and False Negative cases respectively.

During training, we have achieved 100%, 0%, and 0% accuracy, false positive, and false negative respectively. But, during testing, we have achieved 93%, 4%, and 3% in accuracy, false positive, and false negative respectively. The comparison of our proposed method with existing COVID-Net [15] and DCNN [8] is presented in the form of a bar graph which is shown in Fig. 8. From the result it is apparent that our DEEP-CNN based proposed system is able to detect the presence of COVID-19 with 93% accuracy with very less false positive and false negative rates. In this experiment both COVID-Net & DCNN has achieved a very less acuuracy compared to proposed method since they have directly applied the classification without performing any noise removal and segmentation. The experimental results proves the essence of the proposed method by outperforming the other two.

## 6. Conclusion

Early and quick detection of COVID-19 infection is a complex task which is important to reduce the spread of this disease. In this paper, we have presented a Deep CNN architecture to classify normal and COVID-19 patients by using the chest X-ray images. Performance evaluation shows that the proposed method gives a better result compared to other existing methods with about 93% accuracy. We are confident that the findings of this research will help the doctors and other medical practitioners to make accurate decision within a short time. The proposed system could not be tested in extensive environment as very limited number of COVID-19 X-ray images are available till date. Even though CT scan images might have yielded more accuracy, such experiments could not be conducted as COVID-19 CT scan image dataset is not yet available publicly.

## Data Availability

I have referred all the sources of data that we have used in this experiment.

## Compliance with Ethical Standards

1. Conflict of Interest The authors Kishore Medhi, Md. Jamil, and Md. Iftekhar Hussain declare that there is no conflict of interest directly related to the submitted work.
2. Ethical Approval This article does not contain any study with human participants or animals performed by any of the authors. To perform this experiment, we have used the dataset collected from Kaggle which is referred in the dataset section of this manuscript.

---

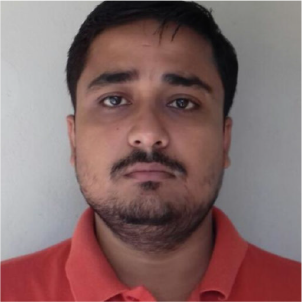

Kishore Medhi was born in Assam, India. He received the B.Tech. and M.Tech. degrees in Information Technology from North-Eastern Hill University, Shillong, India in 2014 and 2017 respectively, where he is currently pursuing the Ph.D. degree at the Department of Information Technology, School of Technology. His current research interests include Machine Learning, Data Mining, Internet of Things, and Image Processing.

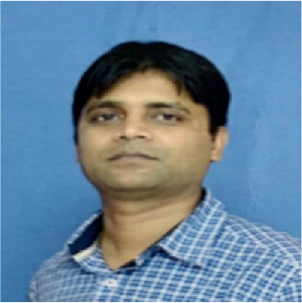

Md. Jamil is an Assistant Professor in the department of General Medicine, North Eastern Indira Gandhi Regional Institute of Health & Medical Sciences, Shillong. His current research includes Tropical diseases.

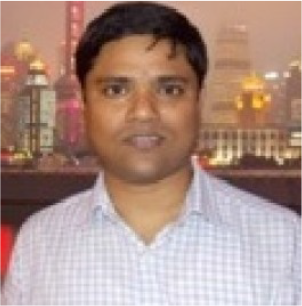

Md. Iftekhar Hussain received his B.E. degree in Computer Science and Engineering from Dibrugarh University, Dibrugarh, India, and M.Tech. degree in Information Technology and Ph.D degree in Computer Science and Engineering from Tezpur University, Tezpur, India. He is an Associate Professor with North-Eastern Hill University, Shillong, India. His current research interest includes Wireless Mesh Networks and Internet of Things. Dr. Hussain is a Life Member of the Indian Science Congress Association and a member of IEEE.

